# Approaches for Interchannel EEG Analysis

**DOI:** 10.1101/2025.02.27.25323027

**Authors:** Wim van Drongelen, Douglas R. Nordli, Mohamed Taha

## Abstract

Here, we present a novel analysis pipeline for scalp EEG signals. With our approach, we focus on the interchannel relationships using standard correlation techniques in the spatiotemporal and frequency domains as well as a recently developed triple correlation-based analysis. Since we compute correlations in multichannel EEG data, we employ (non-standard) bipolar montages with non-overlapping electrode positions to avoid montage-induced correlations in space-, time- and frequency domains. In addition to the correlation studies, we determine the EEG frequency components, phase-amplitude coupling and a burst index. The procedures are tested with simulated signals and illustrated with results obtained from a small patient group.

## Introduction

Individual signals in the electroencephalogram (EEG) recorded with macroelectrodes on the scalp each represent the compound electrical activity in the underlying tissue (e.g. Lopes da Silva, 2004; Nunez and Srinivasan, 2006). Since the discovery of EEG rhythms about a century ago by Berger (1929) the EEG has been a topic of many studies and its application has proven to be of clinical value (e.g., Husain, 2023). Quantitative EEG (qEEG) analysis to complement visual interpretation began after application of computer technology. Computations in the time domain (e.g. Hjorth, 1975; Gotman & Gloor, 1976) and in the spectral domain, based on the fast Fourier analysis algorithm (Cooley & Tukey, 1965) became valuable additions to the EEG interpretation toolbox since the 1960s. Initially the quantitative efforts were realized with so-called minicomputers that were not widely available in the clinic. The application of qEEG took off later in the 1980s, when personal computers became widely available. This started the digital EEG (dEEG) era. A very important addition with the dEEG technology was ability to record an EEG epoch and digitally remontage the measurement off line, thereby replacing a lengthy procedure of sequentially recording epochs of pen-and-paper EEG across different montages. Because of the digitally stored EEG, several other spinoff analyses became available such as source localization procedures (e.g., Scherg & von Cramon, 1985; Fender, 1987; Van Veen et al., 1997) and broad band analyses beyond the typical EEG frequency bandwidth. Historically, the upper bound of EEG frequencies was determined by the limitations of moving pens on paper (or a cylinder), albeit that later moving ink-jets (replacing pens) enabled one to observe slightly higher frequencies. Stability limitations of biological amplifiers and limited analog-to-digital converter bit-depths required the removal of low frequencies at the amplifier input stage. Therefore, the traditional EEG bandwidth from 0.1-1Hz up to 30-70Hz, was predominantly determined by the technology used for recording the signal. With the application of dEEG, these traditional bandwidth limitations widened considerably. Moving pens or ink-jets writing signals on paper were replaced by faster computer displays, A/D converters with greater bit-depth were developed, and cost to store digital EEG data was reduced. These developments all relaxed the technical limitations on bandwidth and an additional interest arose for the lower- and higher frequency components in the EEG recording (e.g. Eissa et al., 2016; Kobayashi, 2021; Lee et al., 2020, 2022).

In spite of the above summarized significant progress in EEG analyses and interpretation, fundamental insight into underlying neuronal processes is still limited, especially how different areas interact in healthy as well as in pathological brain. There are multiple reasons for this, but a critical part of this limited understanding is due to the lack of simultaneous recording of activity of single neuron ensembles and associated EEG, while another essential missing component is a systematic multichannel EEG analysis across well-defined conditions. The purpose of this paper is to evaluate the latter component by combining existing and novel methods to study relationships in multichannel records (van Drongelen, 2018; Deshpande et al., 2023). While this is a first step, our ultimate goal (which is beyond the scope of this paper) is to apply this approach to well-defined epilepsies. Here, as a first step, we demonstrate the robustness and feasibility of our approach by using test signals and illustrating the analysis with a small group of patients.

## Methods, Test Results, and Application to EEG

### Patients

Recordings were obtained in the epilepsy monitoring room at the University of Chicago as part of the clinical evaluation. Consent was obtained from the patients’ legal guardians prior to data collection, and participation was approved by the University of Chicago IRB. Between 2019 and 2024, four patients (3 female, 1 male; 4-12 years of age) were diagnosed and treated for febrile infection-related epilepsy syndrome (FIRES) at our institution. Data from this patient group was used to test all procedures. A selection of these results is illustrated in Figs. 1, 3, 4, 5, and 6.

**Figure 1:**
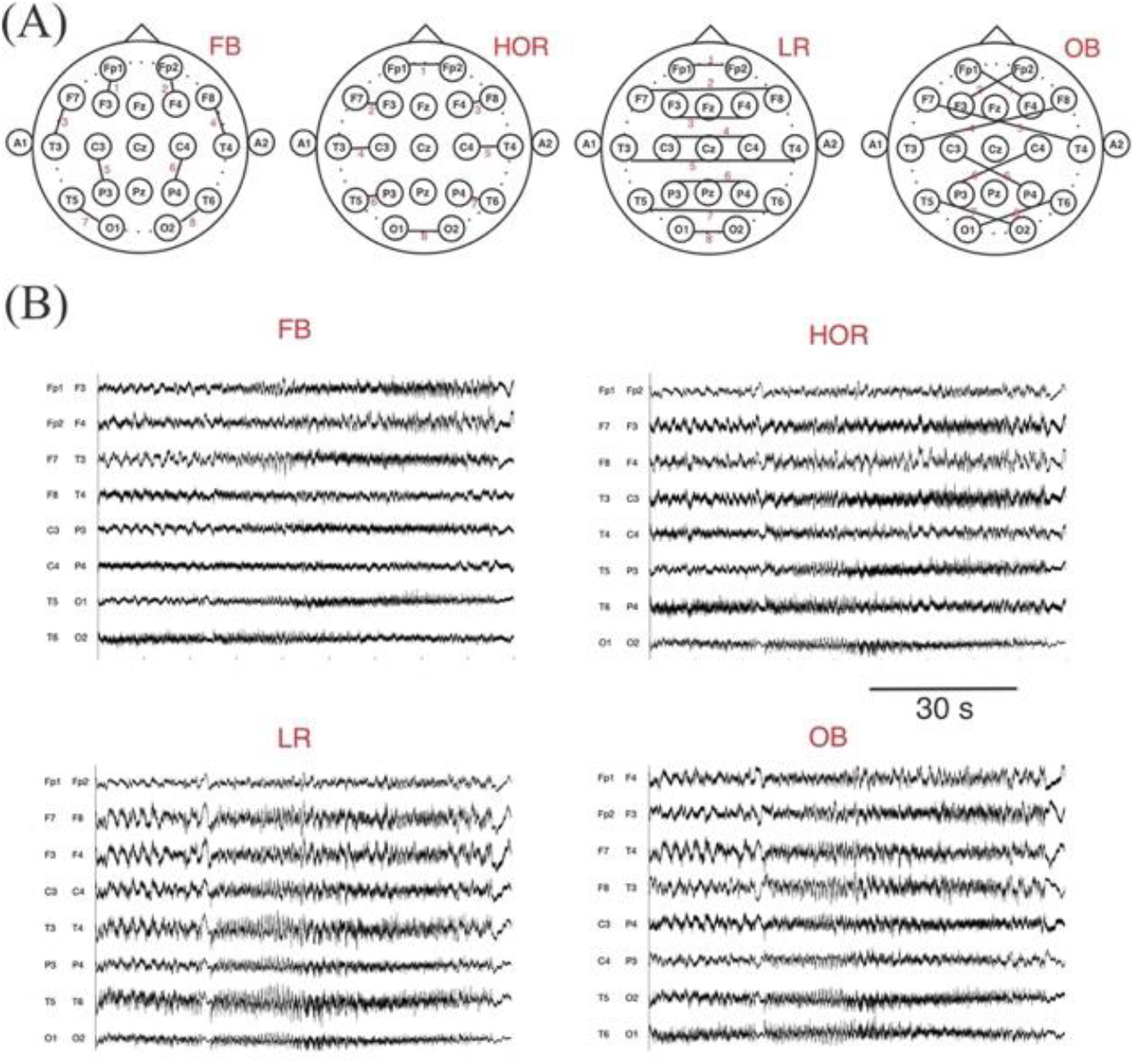
Montage and Example of EEG. A) Bipolar Montages <u>without</u> overlapping electrode positions (lines and channel numbers 1-8) across the eight channels. B) Example EEG (90s) for the eight channels for montages depicted in panel A. Signals are demeaned and divided by their standard deviation.

**Figure 2:**
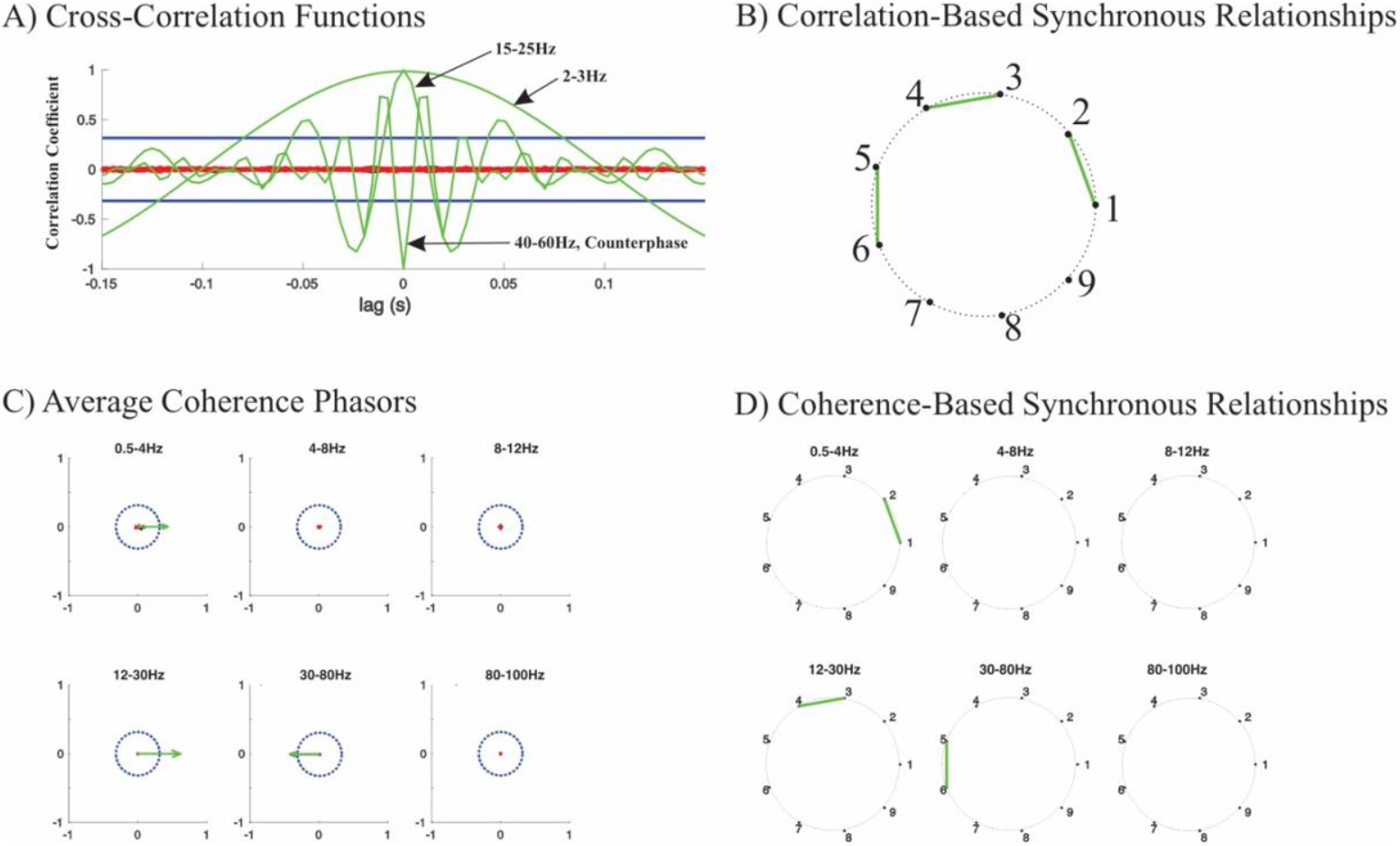
Illustration of the Cross-Correlation and Coherence Analyses Using Test Signals. A) Detail of cross-correlation functions for the test signals with GWN and sine waves in three electrode pairs: Channels 1-2: 2-3Hz; Channels 3-4: 15-25Hz, Channels 5-6: 40-60Hz, with the latter sinusoids in counterphase. The data below the threshold is shown in black (hard to distinguish from the shuffled data) and those exceeding the threshold at some lag(s) as green traces, the shuffled signals are shown as red traces. The threshold is depicted as the blue horizontal lines. The green traces with correlograms exceeding the threshold at lags<10ms were the basis for the diagram in panel B). B) Synchronous relationships in the signals (bandwidth: 0.5-85Hz) based on the cross-correlations depicted in panel A). The green lines represent relationships for correlograms exceeding the correlation threshold (here r > 0.32) at lags<10ms. In this simulation the connectivity diagram shows connections between nodes 1-2, 3-4, and 5-6. The other signals in this simulation (channels 7-9) contained GWN only and (as expected) do not show significant correlations. C) Coherence analysis for the same scenario as in panel A). The average coherence was determined for frequency bands 0.5-4Hz, 4-8Hz, 8-12Hz, 12-30Hz, 30-80Hz, and >80Hz. Each interchannel relationship is shown as one phasor where the magnitude represents the magnitude of the coherence and the angle its phase. The phasors for the EEG data are shown in black, or green if the coherence value exceeds the threshold and its phase deviates <10 degrees from 0 or 180 degrees. The phasors for the shuffled data are shown in red. The green coherence phasors were used to draw the diagrams in D). D) Synchronous relationships in the signals (bandwidth: 0.5-85Hz) based on the coherence analysis depicted in panel C). In contrast to the relationships in panel B), in this analysis connectivity is frequency dependent. Connection between channels 1-2 (at 2-3Hz) shows up in the delta (0.5-4Hz) frequency band, between channels 3-4 (at 15-25Hz) shows up in the beta (12-30Hz) frequency band, and between channels 5-6 (at 40-60Hz, in this example in counterphase) shows up in the gamma (30-80Hz) frequency band, the other channels with GWN only (channels 7-9) do not show any connectivity.

**Figure 3:**
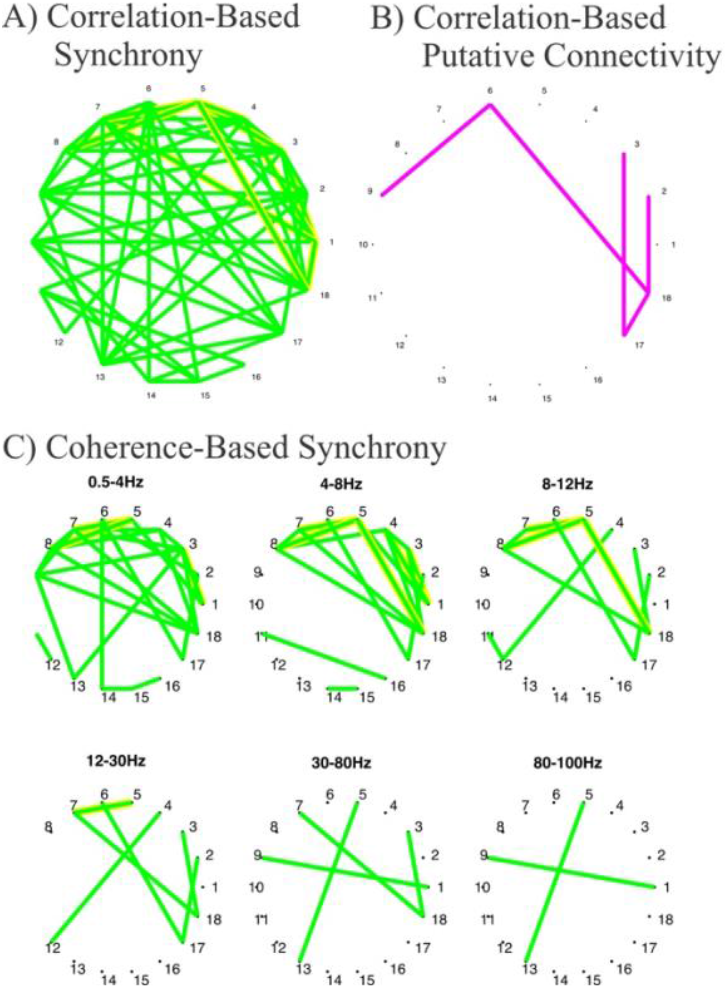
ICross-Correlation and Coherence Analyses Using EEG Signals. A) Same analysis as in Fig. 2B. Here, significant negative correlations are indicated by a yellow border around the green lines. B) Channel relationships in the signals (bandwidth: 0.5-85Hz) based on the cross-correlations. The magenta lines represent relationships for correlograms exceeding the correlation threshold (here r > 0.32) at lags>10ms. C) Same as in Fig 2D. Significant out-of-phase coherence is indicated by a yellow border around the green lines.

**Figure 4:**
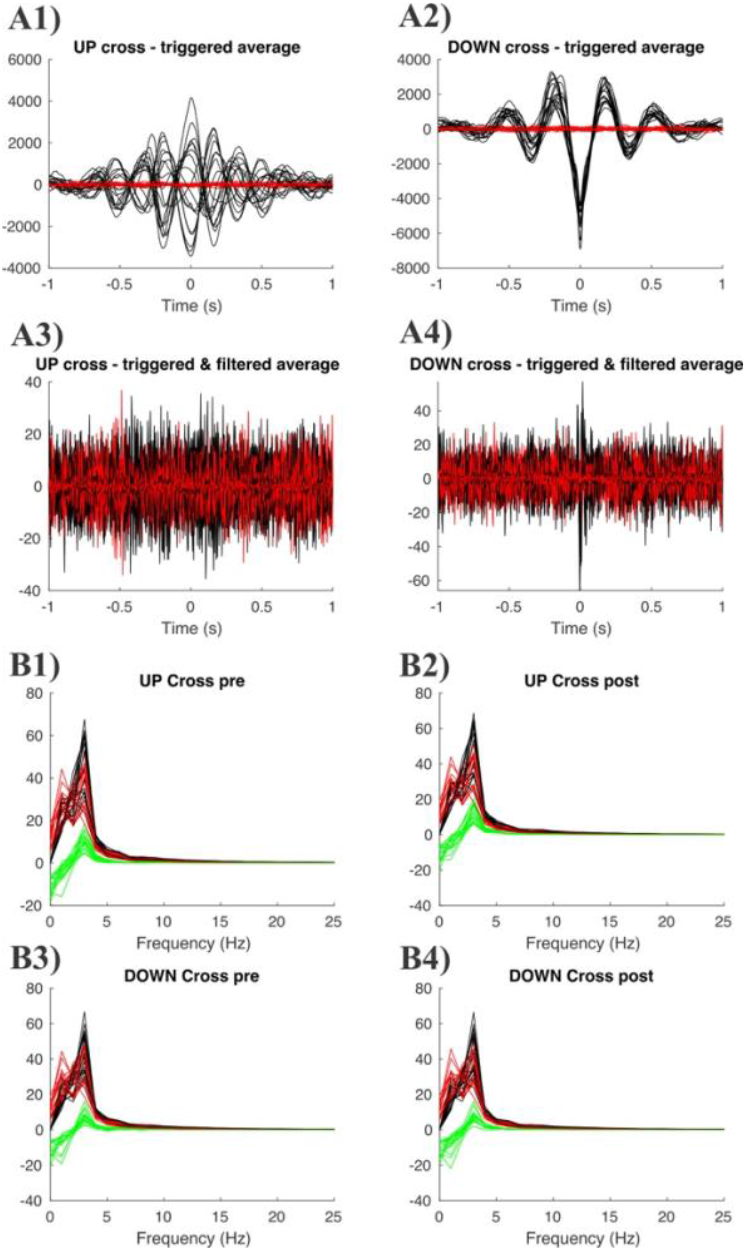
PAC Results Using EEG Based on *fc* = 2Hz. A) The results obtained from averaging time domain using the upward (A1,3) or downward (A2,4) zero-crossing of the average dominant frequency *fc* as the averaging trigger. The results are depicted for broadband (0.5-85Hz) and filtered (40-80Hz) signals. B) Similar as in A), but based on the averages of the power spectra of the broadband EEG, using the upward (B1,2) or downward (B3,4) zero-crossing of the average dominant frequency *fc* as the averaging trigger. Results for the pre- and post-trigger 1s epochs. Note: While this calculation is performed for the broadband signal, in this example the spectra are shown for 0-25Hz only. In all panels black – data; red – shuffled data and in panels B) green is their difference. Prior to analysis, all EEG signals passed a 50/60Hz notch filter.

**Figure 5:**
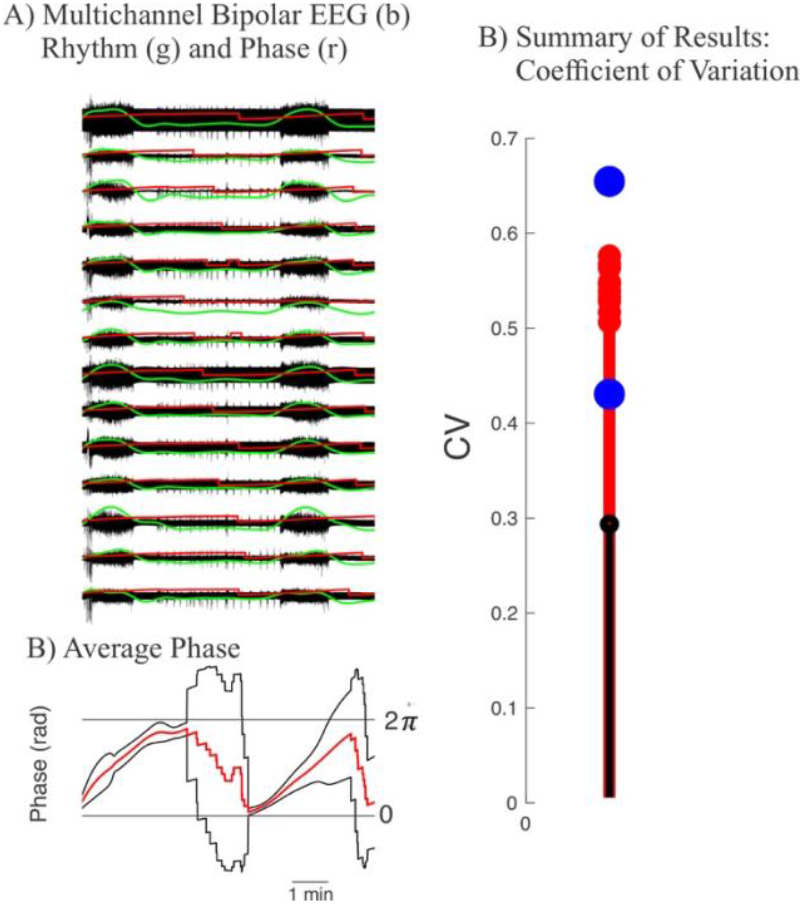
Example of Burst Analysis Applied to EEG. A) 10 min of EEG (black), the rhythm of the super-bursts in each channel (green) and the phase of that rhythm in each channel (red). B) The average phase of the EEG channels (red) and 2 x standard deviation intervals (black). The horizontal lines indicate the 0 and 2*π* limits. C) The results comparing the coefficient of variation of the phase plots of the EEG (black stem) with the same for 10 shuffled data sets (red stems). The blue dots indicate a 5 x standard deviation limit based on the shuffled results.

**Figure 6:**
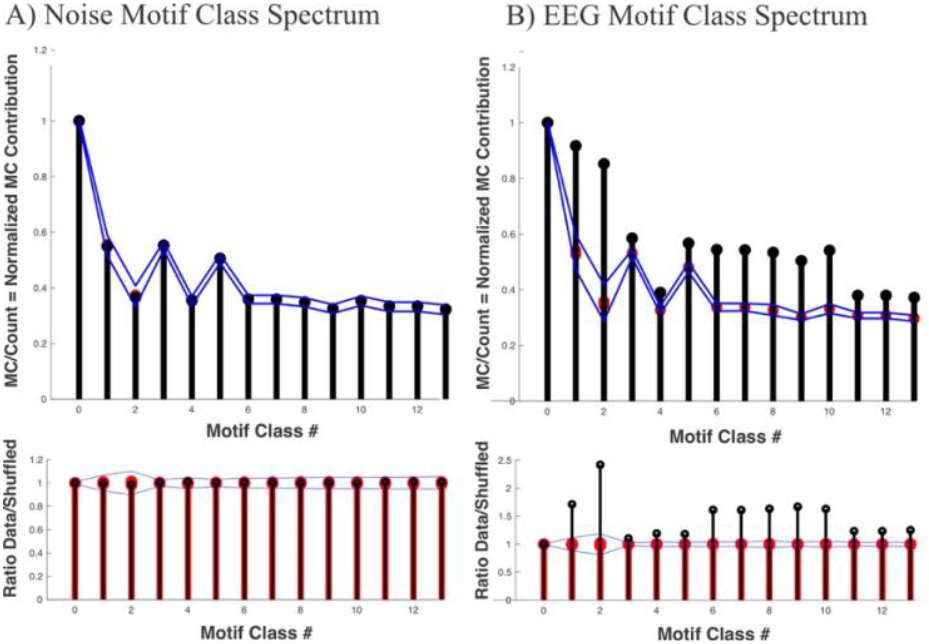
Triple Correlation Analysis on Gaussian White Noise (GWN) using a maximum lag of ±8 samples. A) Top: Motif Class Spectrum for motif classes 0-13 derived from the triple correlation analysis. Contributions from GWN data (black), 25 shuffled data instances (red) and upper and lower thresholds (blue lines; in this example thresholds were ±10 standard deviations). Bottom: Motif-class-ratios for motif classes 0-13. Ratios are data contributions (black, values shown in top panel) and the contributions of the individual shuffled data sets (red, values shown in top panel), relative to the average of the shuffled data. For the shuffled data the ratio was between individual shuffled sets and the remainder. Blue lines indicate upper and lower thresholds (±10 standard deviations). Note that in both panels, the GWN data results (black) are (as expected) located within the thresholds (blue lines) determined by the shuffled versions of the data (red). B) Same as in A) applied to EEG data.

### EEG

EEG data were recorded with a bandwidth of 0.1–85 Hz, sampled at 256 samples/second/channel (Xltek, Natus, Middleton, WI, USA). A representative EEG epoch (≥60s), with minimal eye movement and muscle artifact, was selected by an epileptologist and stored as a segment in the European Data Format (.edf files) for subsequent analysis. The data were processed in Matlab (Matlab, Natick, MA, USA), where raw signals (referenced using a common average) were visually inspected for signal quality. Signals with significant artifacts were excluded, and the final selections were saved as .mat files for further analysis.

### Signal Analysis

Signals were analyzed with in house developed Matlab scripts (For Matlab scripts pls. contact Wim van Drongelen via - in a later stage these will become available on GitHub). We <u>focus our analysis on the quantification of channel interrelationships</u>. Data was evaluated and remontaged in a bipolar fashion where electrodes only contributed **once** to the EEG record to be analyzed (Fig. 1). This procedure was followed to avoid detection of erroneous channel interrelationships due to a common signal. For all analyses, except the triple correlation, the data was filtered with a band-filter (0.5-100Hz), followed by a baseline correction, and (optional) scaling by dividing by the second order correction. For the correction factor we divided all data in the baseline corrected timeseries *x* by 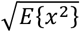.

For a given signal generator, locations of the EEG electrodes relative to that generator is one important factor that determines the polarity of the EEG. In the bipolar approach we used, the polarity can be considered arbitrary. For the cross-correlation-based definition of synchrony, we therefore count positive as well as negative correlations exceeding a threshold. Similarly, in the coherence-based synchrony metric we use phases around 0 degrees (‘in phase’) as well as around 180 degrees (‘counter phase’) for coherence magnitudes above a threshold. In our plots we distinguish between synchrony based on negative correlation and counter phase by a yellow border in the diagrams.

In addition to our evaluation using EEG data, procedures were tested with simulated measurements (Fig. 2). We employed two types of simulations, one consisting of nine signals with Gaussian White Noise (GWN) (created with the Matlab *randn* command), and one test consisting of three GWN signals plus three channel pairs with GWN with superimposed sine waves of different frequencies in each pair:

- Channels 1 and 2 at frequencies 2-3Hz,
- Channels 3 and 4 at frequencies 15-25Hz, and
- Channels 5 and 6 at frequencies 40-60Hz; the 40-60Hz frequencies are in counterphase in the channel pair).

Details of signal averaging, spectral analysis, filters, cross-correlation and coherence are described in van Drongelen (2018) and of the triple correlation in Deshpande et al. (2023). The following is a brief description of the applied procedures.

The bipolar EEG was filtered with a second order Butterworth band filter (0.5-100Hz). Note that the upper value of this band filter didn’t have a major effect on the EEG examples presented here since the data was measured at a bandwidth of 0.1–85 Hz. Data was inspected visually to confirm data quality. All subsequent analyses were performed on the bipolar, filtered EEG data and shuffled versions thereof. For *n* bipolar EEG or shuffled data channels, we computed *n* power spectra, *n*(*n* − 1)/2 cross-correlations, and *n*(*n* − 1)/2 coherence signals (because we studied interchannel relationships, autocorrelation and auto-coherence were not considered and we didn’t consider directionality, thus correlation between *x* and *y* contained the same information as correlation between *y* and *x*). The triple correlation scaled with *n*^3^, because we examined the third order relationships across all channels.

*<u>Power Spectra</u>* (Matlab *fft* command for computing the Fourier transform) were computed on the basis of twelve non-overlapping subsequent 5s epochs (1280 samples) of the 60s bipolar EEG data. These twelve spectra were averaged and depicted in the range 0-25Hz for visual inspection. The same analysis was applied to the shuffled data and the average spectrum was depicted superimposed on the EEG based power spectra.

*<u>Cross-Correlation</u>* measures the correlation between two different signals as a function of the time lag applied to one of the signals. We determined correlations between each pair of signals in the bipolar EEG (bandwidth (0.5-85Hz). We used the Matlab *xcov* command for computing the cross correlation of the demeaned signals. The cross-correlation was determined as the average cross-correlation of twelve non-overlapping subsequent 5s epochs (1280 samples). The cross-correlation result at zero lag is sensitive to large amplitudes in the signals. Therefore, to avoid spurious results we removed deflections beyond 3 standard deviations prior to the cross-correlation procedure. Correlograms were normalized [-1 : 1]. In each of the *n*(*n* − 1)/2 averaged correlograms (Fig. 2A) we determined the largest absolute correlation value and its associated lag. The same procedure was applied to the shuffled data. We produced the following visualizations with these correlation parameters.

(1) A scatterplot of the maximum absolute correlation coefficients versus their associated lags. To minimize the risk for spurious results, only lag values in between -100 and 100ms are examined for further evaluation.
(2) Two network diagrams with the bipolar pairs as nodes in the diagrams. Node pairs were connected with lines if the maximum absolute correlation value between them was above the threshold value (as outlined in Statistics below). One diagram depicted (near) synchronous relationships (e.g., lag < 10ms), (Figs. 2B, 3A). In a similar manner, a similar diagram depicted internode relationships with a lag >10ms (Fig. 3B).

*<u>Coherence</u>* signals (Matlab *mscohere* and *cpsd* commands for computing coherence magnitude and associated phase) were computed across the full bandwidth of the data (0.5-85Hz). We also evaluated in the typical EEG bands: delta (0.5-4Hz), theta (4-8Hz), alpha (8-12Hz), beta (12-30Hz), gamma (30-80Hz), and the remainder, we called high-gamma (80Hz up to the upper limit of the bandwidth). Note that for the sample rate we employed here (256 samples/s) and the associated antialiasing/lowpass filter setting of the EEG recording device, the high-gamma remainder is not important. The coherence was computed in 30 subsequent 2s epochs (512 samples). These calculations were visualized as follows.

(1) A scatterplot of the maximum coherence value against its associated phase. The values of the shuffled data are depicted in the same plots for comparison.
(2) Polar plots of the average complex coherence values (coherence magnitude and phase) in the Beta, Theta, Alpha, Beta, Gamma, and High-Gamma frequency bands (Figs. 2C).
(3) Network Diagrams with the bipolar pairs as nodes in the diagrams. Node pairs were connected if the average coherence magnitude exceeded the threshold (outlined in Statistics below) while the phase angle was below a preset threshold around a 0- or 180-degree phase to represent (near) synchronous relationships (Fig. 2D, 3C).

*<u>Phase Amplitude Coupling (PAC)</u>* was used to assess the relationship between the phase of the low frequency component and the amplitude of the higher frequency oscillations. The average frequency of the maxima in the power spectra across the bipolar channels in the EEG signal was determined. This frequency (*fc*) was used to filter the EEG using a zero-phase FIR band filter (order: 10; bandwidth: 0.5 × *fc* – 1.5 × *fc* Hz) resulting in a low frequency component of the channels. Two trigger options were explored:

(1) The zero-crossings of the low frequency filtered EEG.
(2) The transitions in the phase signal obtained from the Hilbert transform of the low frequency filtered EEG (used unless stated otherwise).

We determined two trigger types: upward and downward zero-crossings. Based on these triggers:

(a) We averaged the broadband time domain signal. The time domain average produces a timing sensitive signal indicating the signals around the zero crossings. These signals were filtered (FIR, order: 10, bandwidth: 40-80Hz) to detect high frequency components around the triggers.
(b) We determined the power spectra 1s before and 1s after the trigger. The power spectra indicated presence of frequency components in the second before and after the trigger. In contrast to the time domain averages where results depend on the phase of the oscillatory signals across the individual trials, the power spectra only reflect the presence of such signals, whether in- or out of phase.

As control we randomly selected the same number of epochs from the EEG and performed the same time and frequency domain analyses as in (a) and (b) above. A typical result is shown in Fig. 4.

*<u>Burst Analysis</u>* was applied in records that demonstrated repetitive groups of bursting (super-bursts) across multiple channels (Fig. 5A). In step 1 of this analysis, the peak frequency of the intraburst activity was determined followed by the application of a narrow band filter around this peak frequency. In steps 2 and 3, the output of the filter was rectified and passed through a leaky integrator (e.g. Suresh et al., 2016) (green lines Fig. 5A). This result that showed a signal that represented presence of the super-bursts was used as input to the Hilbert transform to determine the phase of the super-burst across channels (red lines, Fig. 5A), thereby giving a visual impression of the relationships across the EEG record. The application to a multichannel recording shows the level of synchronous super-burst activity and the average of the phase plot gives an impression how synchronous the EEG is at the temporal scale of the super bursts (Fig. 5B).

In addition, the coefficient of variation of the phase plots across channels is a metric for the super-burst synchrony. We determine significance of this metric by using the comparison of the same metric of shuffled data (fig. 5C) – shuffling was done over both time and space.

*<u>Triple Correlation</u>* measures the simultaneous interaction between three different variables. The following is based on the triple correlation uniqueness (TCU) theorem and its application for analysis of neuroscience data (Deshpande et al., 2023). For the triple correlation analysis, we also employed the assumption that deflection of the EEG signal from the 0-line reflects the important component of the signal. Therefore, we use full-wave rectified (absolute value) EEG for the determination of the triple correlation. Prior to <u>triple correlation analysis</u>, we preprocessed the EEG by a rather narrow band-filter (1-35Hz), followed by a baseline correction, full-wave rectification and scaling by dividing by the third order correction. For the correction factor we divided all data in timeseries *x* by 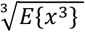 (Appendix II, Fig. II,2), where *E*{*x*^3^}, is the Expectation of *x*^3^ of the samples in the timeseries. The Expectation was estimated as the average of *x*^3^ in each channel. This procedure was employed because in triple products the relationship between positive and negative contributions to the correlation result is not as easily interpreted as e.g. in cross-correlation. Triple correlation contributions were computed as a function of the two spatial and two temporal lags. This resulted in a 4-dimensional correlation matrix (*C*3) and an associated 4-dimensional matrix (*No*) in which we kept track of the number of contributions for each lag combination. Dividing sum of the contributions by the number of contributions (*C*3(*n*1, *t*1, *n*2, *t*2)/*No*(*n*1, *t*1, *n*2, *t*2)) produced a 4-dimensional matrix with the mean contribution (mu) for each lag contribution. This mu-matrix was used to compute the Motif-Class Spectrum, with 14 motif classes (Deshpande et al., 2023) (Appendix I), that governed the underlying EEG. For each motif class we also determined the number of contributions in that class (count).

To obtain control data sets, the bipolar data was randomly shuffled across time and across channels to remove temporal and interchannel relationships. The same preprocessing steps used for the data were also used for the shuffled data. Statistics were determined with the shuffled data; we used 25-100 instances of shuffled control data. Using the third order normalization described above, 3^rd^ order activity levels (represented by Motif Class 0 values) for the EEG and shuffled EEG data versions are kept identical (Fig. 6). The motivation for this approach is that we want (1) to interpret motif class results independent of the activity level and (2) to include the effects of the pre-processing in the statistics of the control data. Especially including the effect of filters is essential because the so-called raw EEG is recorded with a bandpass filter and random shuffling destroys the filter effect. The stem plots in Fig. 6A show an example of metrics derived Motif Classes 0-XIII applied to a multichannel random noise signal. The same for an EEG epoch is depicted in Fig. 6B.

## Statistics

To determine statistical thresholds we employed Chebyshev’s theorem, applicable for any distribution with a given mean and finite variance: i.e., the fraction of data **at least** within ±k × *std* (*std* - standard deviation) range is equal to 1 − 1/k^2^(Tchebichef, 1867). At a 1% significance level that results in ±k = 10. Note that this value exceeds the so-called Rose criterion stating a significant signal must be at least outside a range of ±5 × *std* against a control/background (Bushberg, et al., 2006; Rose, 1973).

In many cases a highly significant correlation coefficient was found, but sometimes at rather small correlation levels. Therefore, in addition, we only considered significant correlation coefficients accounting for at least 10% of the variance (i.e., correlation coefficient > 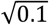). The combined approach demanding at least statistical significance of 1% and at least a correlation of 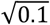, generated a conservative threshold for signal versus noise separation. For the *<u>PAC analyses</u>*, where we looked into the events associated with low frequency component phase, the controls were composed of signals around randomly positioned triggers. *<u>Correlation</u>* and *<u>Coherence</u>* were interpreted by comparison of the result for the bipolar data with their spatiotemporal shuffled version. The statistics for the shuffled data determined the threshold as described above.

*<u>Burst Analysis</u>* results are visually evaluated using the average phase of all channels, and we use the coefficient of variation (CV) (standard deviation divided by the mean) of the phase results for statistical evaluation. The CV of the data and mean ± 5 × standard deviation of spatiotemporal shuffled versions thereof provide a significance criterion. In the example in Fig. 5 we used 10 shuffled data sets.

*<u>Triple correlation</u>* based motif classes are based on averages and according to the central limit theorem these follow a normal distribution. If we apply a Bonferroni correction of comparing 14 MCs, using a 1% significance level, we should demand a level of 0.07%. The 10 standard deviation criterion we use is far beyond that.

The mean and std old the MCs for the shuffled data is OK.

You might consider the std as an SEM. This means that the mean that’s not depending on the number of contributions (size of EEG and max lags) while the std/SEM does. It’s in essence an averaging procedure where the estimate of the MCs becomes more reliable with increased N.

## Discussion

In this work we describe the steps included in an analysis pipeline we developed for the evaluation of interchannel relationships in EEG data. We test the procedure and demonstrate the use of correlation analysis to detect synchronous activities in the time and frequency domains (Figs. 2, 3). We carefully avoid detection of spurious correlation, using montages with non-overlapping electrode positions (Fig. 1). Especially in scalp EEG where electrodes may have equally important signal contributions, correlation between channels can easily be introduced by a common reference or a common electrode. Even with the so-called Laplacian montage (Hjorth, 1975), one might introduce significant correlations because a set of montaged EEG signals may share a (weighted) contribution of a common electrode position. While we avoid common electrode positions across bipolar montages we use, we do find strong and significant proof of synchrony across brain regions (Fig. 3). In this context, it is important to note that similar problems with intracranial EEG can be enormously reduced by using an extracranial location for the reference electrode – that is because the intracranial signals are usually orders of magnitude stronger than the extracranial ones so that an extracranial common component does not introduce a strong (false) interchannel relationship between intracranial data.

In our network analysis, we employed the triple correlation estimate that was recently introduced for application in neuroscience (Deshpande et al., 2023). The approach is based on the triple correlation uniqueness (TCU) theorem (Yellott & Iverson,1992; Yellott, 1993; Victor, 1994), stating that the triple correlation (in contrast to the widely applied cross-correlation and auto correlation) provides a unique representation of the underlying data, in our case the neuronal activity pattern. Deshpande et al., (2023) applied this to characterize spike activity in the form of spike rasters, and to quantify network motifs. These motifs were grouped in fourteen motif classes (MC), numbered 0-XIII (Appendix – Fig. I,1). Here, we adjusted the same approach to EEG, a continuous signal. To make this work, several critical adjustments were made. First, we rectified the signal, recognizing that signal amplitude rather than its polarity is important. This also addresses a more technical aspect, namely that the Expectation of the third order moment of zero-mean noise is zero (e.g., Lohman &Wirnitzer, 1984), which would lead to impractical small values of the EEG’s triple correlation. Second, for statistical evaluation, we compare the outcome of the triple correlation analysis to the outcomes from shuffled versions subjected to identical pre-processing steps. Third, to make sure that we compare higher order motif classes in the data versus its shuffled versions, we set the activity level (MC 0) to be equal in data and shuffled versions (similar to the approach used for spike rasters) by dividing the pre-processed data by its third moment. These steps result in a motif class spectrum that allows for interpretation of motifs that govern the underlying EEG (Fig. 6). The steps we employed for the triple correlation analysis reflect the same principles we used in the cross-correlation and coherence analyses. Also, in these analyses we did not distinguish the polarity of the EEG, and we also normalized (by the second moment in these cases) to avoid effects of amplitude differences across data and shuffled versions thereof.

The idea behind the statistical analysis we employ is that we only want to consider highly significant effects. Therefore, we use Chebyshev’s theorem to determine the statistical threshold of ten standard deviations, one that is not just valid for normally distributed data but for any type of data instead (Tchebichef, 1867). In addition, we use an overarching condition to only consider correlations across signals in multichannel EEG above 10% to increases the likelihood that we identify channels with a strong functional connection or a clear synchronization. A reason for our rigorous statistical approach is not just to ignore minor effects, but to increase the probability that effects that cross our threshold allow the evaluation of single records in addition to finding differences across groups.

Based on our tests and preliminary analyses, we conclude that the proposed pipeline for the evaluation of network properties from EEG provides a promising avenue to evaluate patient data. Our laboratory plans to apply this approach to evaluate functional connectivity in pediatric patients with monogenetic epilepsies, and to use this tool for diagnosis and evaluation of therapy.

## Data Availability

All data produced in the present study are available upon reasonable request to the authors

## Acknowledgement

Part of this work was supported by The Chan Zuckerberg Initiative (CZI).

## Appendix I: Overview Motif Classes

The triple correlation analysis presented here is an extension of the approach to characterize neuronal spike activity. Details are described in Deshpande et al. (2023). The diagram summarizing the so-called motif classes, based on three-node motifs is included here for convenience.

**APPENDIX: Figure II,1.**
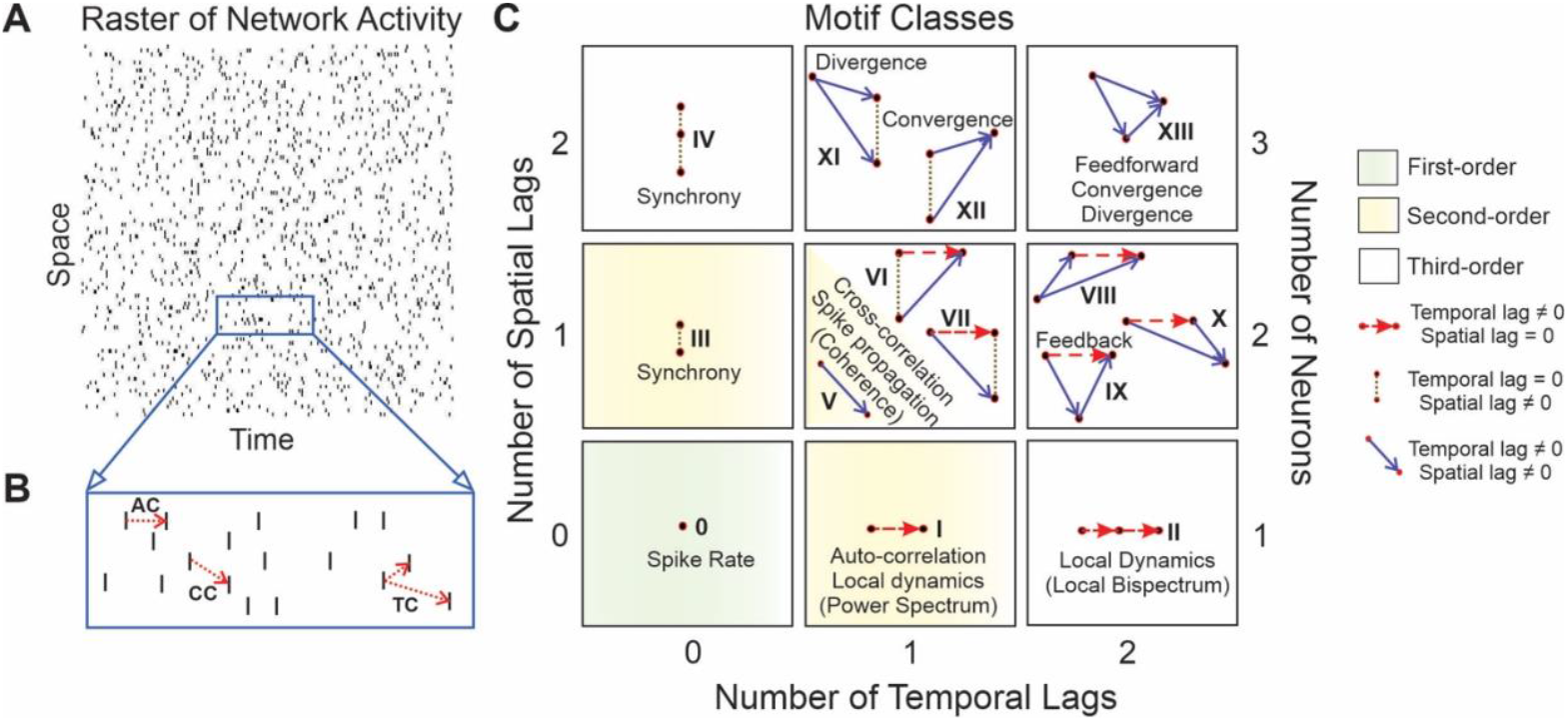
Overview of motif classes extracted from the triple correlation of an activity pattern (from Deshpande et al., 2023). A) An activity pattern represented in a raster. B) Correlation examples. AC – Autocorrealtion; CC – Cross-correlation; TC – triple correlation. C) Motif Classes 0-XIII extracted from the (unique) triple correlation of an activity pattern. The classification is based on the number of spatial and temporal lags of the correlation contributions and the symmetries that that occur with these. Details for the procedure are described in Deshpande et al. (2023). *Figure reprinted with permission from authors and under the terms of the Creative Commons CC BY license from Deshpande et al. (2023)*.

## Appendix II: Scripts

The analysis pipeline employed in this paper consists of the number of Matlab scripts, most included in the PipelineEEG_AutoFunction (Fig. II,1). Two functions outside this pipeline function are (1) a function to read EEG data in edf format and (2) a script that characterizes burst activity in the EEG (Fig. II,1).

Preprocessing is an important aspect of the analysis used here. This includes filtering but also normalization of amplitudes. These operations and the functions in which these are implemented are summarized in Fig. II.2.

For Matlab scripts pls. contact Wim van Drongelen via

**APPENDIX: Figure II,1.**
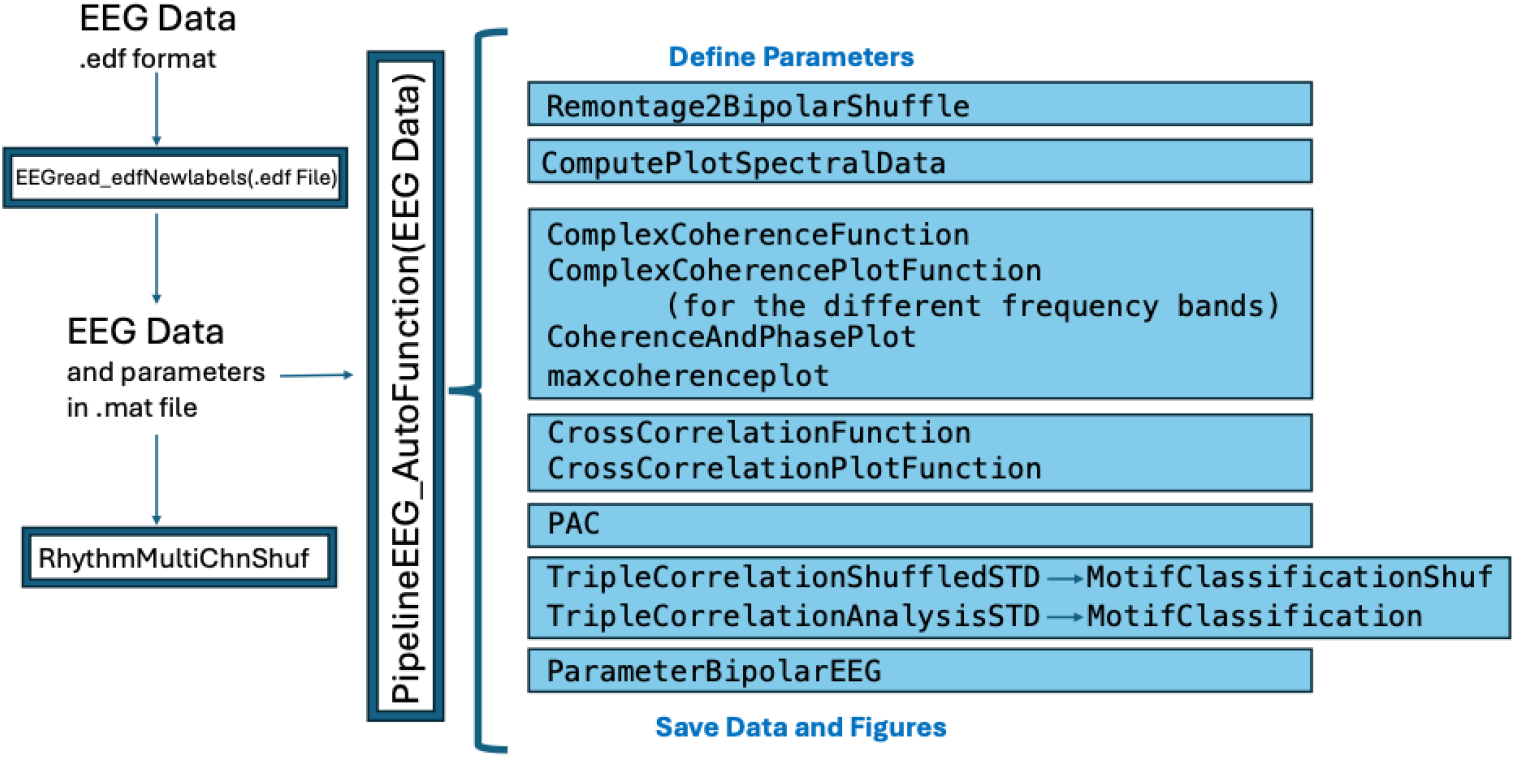
Overview of the Pipeline Functions Applied to the EEG. The pipeline starts with the conversion from edf format into .mat format. The analysis steps are done with functions that are included in the principal function PipelineEEG_AutoFunction that uses the .mat file as its input. This principal function includes the parameter definition and is responsible for saving the resulting data and figures depicting it. The functions in the principal function, listed with the blue background, are performed sequentially from top to bottom. These functions are responsible for the pre-processing (see Fig. II,2), spectral analysis, coherence, cross-correlation, phase-amplitude coupling (PAC) analysis, triple correlation analysis, and the extraction from selected result parameters. The function RhytmMultiChnShuf is separate because it only applies to ictal EEG with rhythmicity.

**APPENDIX: Figure II,2.**
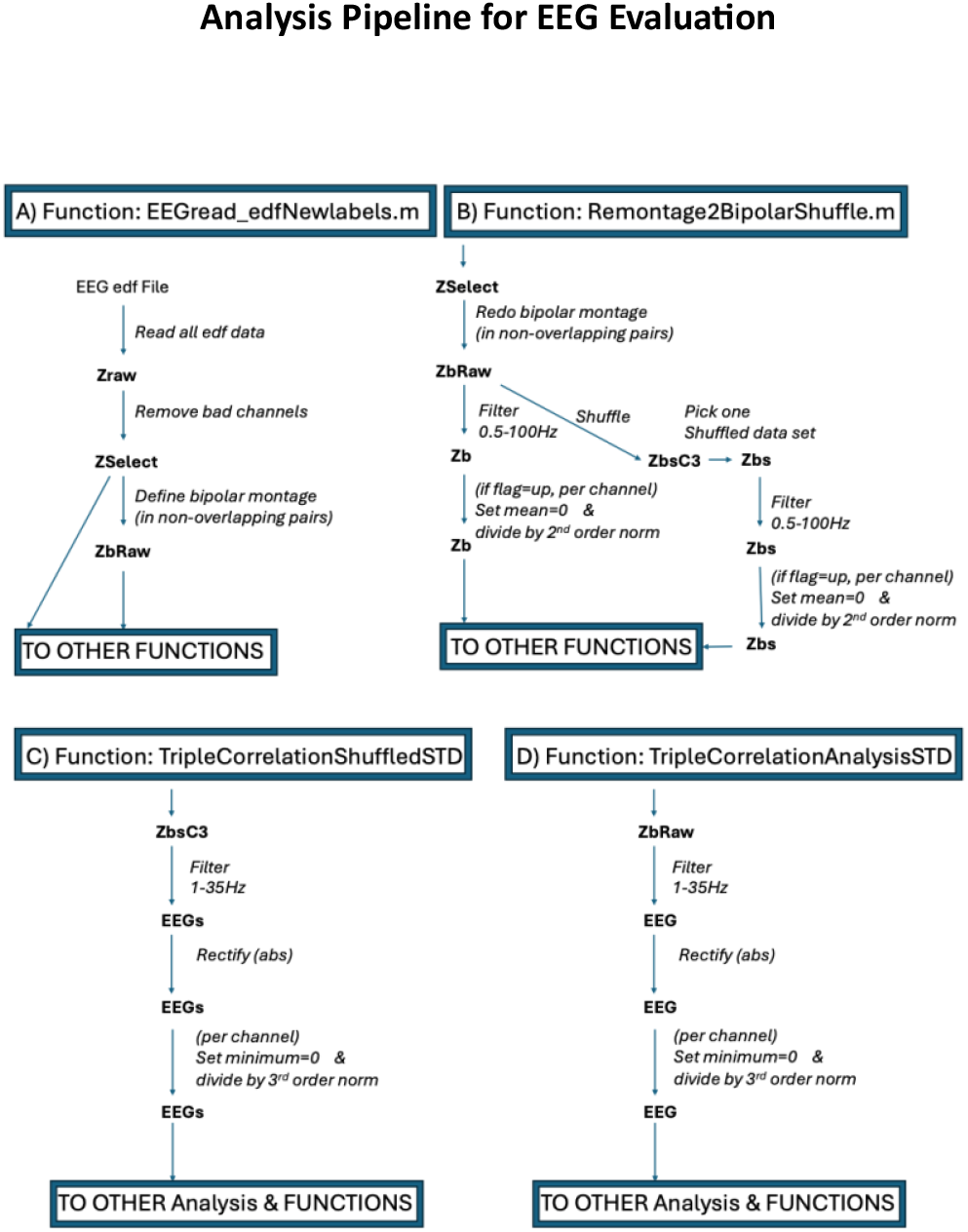
Overview of the Preprocessing Procedures Applied to the EEG. The four functions (A-D) that are involved in the pre-processing of the EEG and the shuffled versions thereof. Functions A-D are listed by their name, variable names are in bold and processing steps in italic. A) The first step is reading the EEG from a clip in edf (European Data Format). After removal of bad channels, the data is remontaged. B) For all subsequent analyses, except the triple correlation analysis, EEG data is filtered from 0.5-100Hz and normalized if the normalization flag is up. The unfiltered data is shuffled for the triple correlation and one shuffled set is used for the other analyses. This single shuffled set is, as the EEG signals, filtered from 0.5-100Hz and normalized if the normalization flag is up. C) Prior to the triple correlation analysis, the unfiltered shuffled data is filtered (1-35 Hz) prior to the triple correlation analysis. Next, the data is rectified, divided by the standard deviation and the minimum value is set to zero. D) Prior to the triple correlation analysis, the unfiltered EEG itself is preprocessed as in C).

